# Analytical validation and clinical utilization of K-4CARE^TM^: a comprehensive genomic profiling assay with personalized MRD detection

**DOI:** 10.1101/2023.11.07.23298196

**Authors:** Thien-Phuc Hoang Nguyen, Tien Anh Nguyen, Nam HB Tran, Van-Anh Nguyen Hoang, Hong Thuy Thi Dao, Vu-Uyen Tran, Yen Nhi Nguyen, Anh Tuan Nguyen, Cam Tu Nguyen Thi, Thanh Thuy Do Thi, Duy Sinh Nguyen, Hoai-Nghia Nguyen, Hoa Giang, Lan N Tu

**Affiliations:** Medical Genetics Institute, Ho Chi Minh city, Vietnam; Gene Solutions, Ho Chi Minh city, Vietnam

**Keywords:** comprehensive genomic profiling, tumor mutational burden, microsatellite instability, minimal residual disease

## Abstract

**Background:** Biomarker testing has gradually become standard of care in precision oncology to help physicians select optimal treatment for patients. Compared to single-gene or small gene panel testing, comprehensive genomic profiling (CGP) has emerged as a more time- and tissue-efficient method. This study demonstrated in-depth analytical validation of K-4CARE, a CGP assay that integrates circulating tumor DNA (ctDNA) tracking for residual cancer surveillance.

**Methods:** The assay utilized a panel of 473 cancer-relevant genes with a total length of 1.7 Mb. Reference standards were used to evaluate limit of detection (LOD), concordance, sensitivity, specificity and precision of the assay to detect single nucleotide variants (SNVs), small insertion/deletions (Indels), gene amplification and fusion, microsatellite instability (MSI) and tumor mutational burden (TMB). The assay was then benchmarked against orthogonal methods using 155 clinical samples from 10 cancer types. In selected cancers, top tumor-derived somatic mutations, as ranked by our proprietary algorithm, were used to detect ctDNA in the plasma.

**Results:** For detection of somatic SNVs and Indels, gene fusion and amplification, the assay had sensitivity of >99%, 94% and >99% respectively, and specificity of >99%. Detection of germline variants also achieved sensitivity and specificity of >99%. For TMB measurement, the correlation coefficient between whole-exome sequencing and our targeted panel was 97%. MSI analysis when benchmarked against polymerase chain reaction method showed sensitivity of 94% and specificity of >99%. The concordance between our assay and the TruSight Oncology 500 assay for detection of somatic variants, TMB and MSI measurement was 100%, 89% and 98% respectively. When CGP-informed mutations were used to personalize ctDNA tracking, the detection rate of ctDNA in liquid biopsy was 79%, and clinical utility in cancer surveillance was demonstrated in 2 case studies.

**Conclusions:** K-4CARE^TM^ assay provides comprehensive and reliable genomic information that fulfills all guideline-based biomarker testing for both targeted therapy and immunotherapy. Integration of ctDNA tracking helps clinicians to further monitor treatment response and ultimately provide well-rounded care to cancer patients.

## Introduction

Gene mutation-targeted therapy was first approved by the U.S Food and Drug Administration (FDA) in 2001 [1]. Many ground-breaking developments since then have revolutionized cancer treatment and transformed the field of cancer diagnosis. Such progress is tightly linked with the advancement in genetic sequencing technologies, which have uncovered the mutational landscapes of cancer and hence revealed new actionable alterations [1]. Once a targeted therapy is approved, it is crucial to identify patients with the right genomic alterations predictive of response to the treatment. Therefore, biomarker testing has been recommended as the standard of care to empower precision oncology. As the number of therapies increases, the list of biomarkers also expands, not just in number but also in the complexity. Moving beyond single genomic alterations, some complex biomarkers have emerged, such as tumor mutational burden (TMB) and microsatellite instability (MSI). For the use of immune checkpoint inhibitors (ICIs) in advanced cancer, TMB and MSI have both been approved by the FDA as independent predictive biomarkers in multiple solid tumors, besides programmed death-ligand 1 (PD-L1) positivity [2].

In real-world practice, biomarker testing is often run as single-gene, hotspot or small panel assays due to their low cost, simple workflow and quick turnaround time. Particularly in developing countries, popular methods are based on polymerase chain reaction (PCR) such as digital droplet PCR to detect single point mutations; or immunohistochemistry (IHC) and fluorescent in situ hybridization to detect gene amplification or rearrangement [3; 4]. Next-generation sequencing (NGS) is increasingly popular but still limited for pre-selected gene panels. The main disadvantage of this practice is the inadequate coverage of all companion biomarkers and failure to analyze complex genomic signatures such as TMB and MSI, which could ultimately lead to patients missing treatment opportunities. For instance, it was reported that for lung carcinoma, the rate of *EGFR* testing was the highest (>80%) while the rates of *ALK* and *ROS1* testing were remarkably lower [3], despite the fact that *ALK* and *ROS1* mutations could occur in up to 10% of the cases [5; 6]. Therefore, comprehensive genomic testing (CGP) that analyzes hundreds of genes in a single assay could be a more efficient solution. CGP consolidates results for multiple biomarkers for both approved and emerging therapies while conserving precious biopsy samples. Knowing all treatment options upfront could help physicians select or change therapy promptly when the disease progresses.

The major drawbacks of CGP are the cost and necessity of qualified equipment and highly skilled labor. Hence, CGP assays are either not available or not affordable for majority of patients in resource-restricted countries. Moreover, the quality of formalin-fixed paraffin-embedded (FFPE) tumor tissues, the primary resource for CGP, remains a big challenge in these countries. DNA and particularly RNA from FFPE could be heavily degraded due to problems in formalin fixation and storage condition. Limitation in both sample quantity and quality thus requires optimization, and standardized CGP protocols developed for FFPE samples of high quality might not be applicable. Furthermore, CGP is criticized to detect many mutations that are not treatment-meaningful [7]. If this mutation profile could be leveraged to guide detection of circulating tumor DNA (ctDNA) in the plasma, this would then facilitate real-time monitoring of tumor burden and patient’s response to treatment. Meaningful clinical benefits of ctDNA monitoring to detect minimal residual disease (MRD) have consistently been documented [8].

In this study, we established K-4CARE^TM^ (K4C), a tissue-based CGP assay utilizing a panel of 473 cancer-associated genes to detect single nucleotide variants (SNVs), small insertion/deletions (Indels), gene amplification and fusion, as well as TMB and MSI assessment. The assay was thoroughly validated using reference standards and optimized for clinical samples of cancer patients in Vietnam. For selected types of cancer, the K4C assay also integrated ctDNA detection in the liquid biopsy for cancer surveillance.

## Materials and Methods

### Sample collection

Genomic DNA from formalin-fixed paraffin-embedded (FFPE) tumor samples and matching white blood cell (WBC) samples of cancer patients were obtained from the Medical Genetics Institute, Ho Chi Minh city, Vietnam. All samples were de-identified and stored for less than 2 years. In total, 282 FFPE samples from more than 10 types of cancer were included in the study. The full list and allocation of samples for different experiments are in Table S1. The list of commercial reference standards used to assess the performance of the K4C assay is in Table S2. For ctDNA detection, plasma samples of 40 patients that had either colorectal cancer (CRC) or breast cancer were used in the analysis (Table S1).

### Library preparation and sequencing for FFPE and WBC

Genomic DNA was already isolated from FFPE and WBC samples as previous described [5; 9; 10; 11]. DNA fragmentation and library preparation for paired FFPE and WBC samples were performed using the NEBNext Ultra II FS DNA library prep kit (New England Biolabs, USA) following the manufacturer’s instructions. Libraries were pooled together and hybridized with predesigned probes for a panel of 473 cancer-relevant genes (Integrated DNA Technologies, USA). This gene panel was curated from large public cancer databases, high-impact cancer genomic studies, and particularly in-house database of prevalent mutations from >10,000 cancer patients. The panel included entire exons of 473 genes, promoter region of *TERT*, and selected introns of 6 common fusion genes (Table S3). Massive parallel sequencing of DNA libraries was performed on the DNBSEQ-G400 sequencer (MGI, China) with an average depth of 500X for FFPE and 500X for WBC samples.

In the experiment to validate TMB, DNA libraries were also hybridized to the xGen Exome Hyb Panel v2 (IDT, USA) that covered the entire human exome. Sequencing was performed on the DNBSEQ-G400 sequencer (MGI, China), with the average depth for FFPE and WBC samples of 250X and 175X, respectively. In the experiments to compare K4C assay with the commercial TruSight Oncology 500 (Illumina, USA), the library preparation and target enrichment for FFPE samples were performed following the manufacturer’s protocol. DNA libraries were sequenced on the NextSeq 550 (Illumina, USA) in paired-end 150-cycle runs, with the average depth of 1000X.

### Bioinformatics analysis

For quality control, we used Illumina DRAGEN^TM^ Bio-IT Platform (v3.10) to trim low-quality bases and adapter sequences in fastq files and to remove duplicated reads from the bam files. FFPE samples that passed quality control (QC) of the K4C assay must have a mean depth greater than or equal to 150X and a percent coverage at 100X exceeding or equal to 75%. The failure rate was 10.4% and samples that failed QC were removed from analysis.

Somatic variants were called from bam files by the DRAGEN^TM^ Somatic pipeline, using matched tumor-normal pairs. To reduce the number of artifacts related to formalin treatment and tissue sample processing, post somatic calling filtering was performed including ignoring soft-clip bases and removing clustered events. VEP (version 105) [12] was used to predict the effects of variants and annotate them against dbSNP [13], ClinVar [14], and COSMIC [15] databases. For gene amplification, DRAGEN^TM^ Somatic CNV Calling pipeline was utilized. Ninety WBC samples were used to build a reference baseline, from which the normalized copy ratio value was calculated. The gain or loss of the targeted genes was called, with bin segment = 100 bp. Gene amplification was detected when the copy ratio of segment mean ≥ 2.0. For gene fusion, the aligned bam files from DRAGEN^TM^ with the marked soft-clipped reads were analyzed using Factera (v1.4.4) [16] with default parameters. Fusions were detected when minimal 2 breakpoint-spanning reads were reported. The OncoKB database [17] (last updated 10/02/2023) was then used to identify actionable alterations eligible for FDA-approved drugs (level 1-2).

Germline variants were called from bam files of WBC samples by the DRAGEN^TM^ Germline pipeline. Variants were then classified according to the guidelines of The American College of Medical Genetics and Genomics (ACMG) as previously described [11; 18]. Pathogenic and likely pathogenic variants were reported.

TMB calculation was performed using our in-house developed script to divide the total number of eligible somatic variants by the size of the interrogated panel. An eligible somatic variant must meet all of the following criteria: (1) pass filtering parameters of the variant calling pipeline, (2) not likely a germline variant as filtered by the dbSNP database [13], (3) locate within the coding region, (4) not a synonymous mutation, (5) have VAF ≥ 5%, allele depth ≥ 5X, total depth ≥ 15X. The panel size was counted for the bases within coding regions with the minimal total depth ≥ 15X. The threshold for TMB-High (TMB-H) was determined as 10 mutations/Mb. For MSI status, unstable microsatellite loci were detected by MSIsensor-pro (v1.2.0) [19] in matched tumor-normal mode. If the proportion of unstable loci among all detected microsatellite loci was at least 20%, the sample was determined as MSI-High (MSI-H).

For samples analyzed using the TruSight Oncology 500 kit, the TSO500 analysis pipeline was executed utilizing the DRAGEN^TM^ TSO500 (v2.1) software on the Illumina DRAGEN^TM^ Bio-IT Platform (v3.10). One sample (1/40) that failed the recommended QC metrics according to the TSO500 manufacturer’s guidelines was excluded from the analysis.

### MSI analysis by PCR

DNA from paired FFPE and WBC samples were subjected to fluorescent multiplex PCR reactions using the MSI Analysis System, Version 1.2 (Promega, USA) to examine 5 mononucleotide repeat markers: BAT-25, BAT-26, MONO-27, NR-21 and NR-24. Samples were run on the SeqStudio^TM^ genetic analyzer (ThermoFisher, USA). Those with at least 2 altered markers were classified as MSI-H, while others were classified as microsatellite stable (MSS).

### Plasma ctDNA detection

All FFPE-derived mutations were ranked using our proprietary scoring algorithm as previous described [9; 10]. The main criteria included VAF in FFPE, predicted pathogenicity and deleteriousness, mutation type, reported frequency in large cancer databases, and internal validation as a tumor-derived mutation. The top ranked mutations specific to each patient were then used for ctDNA tracking in the plasma samples.

cfDNA was already extracted from plasma samples as previously described [9; 10]. The minimum amount of cfDNA required was ≥ 0.15 ng/uL or total of ≥ 3 ng. cfDNA fragments with the selected mutation sites were amplified using KAPA HiFi DNA Polymerase (Roche, USA) and targeted primer pairs (PhuSa Biochem, Vietnam). Amplified cfDNA fragments were sequenced on the NextSeq 2000 system (Illumina, USA) with an average depth of 100,000X per amplicon. Amplicons with less than 10,000X coverage were considered unsuccessful. Variant calling was performed using mpileup from Samtools (v1.11) as previously described [9; 10]. A plasma sample that had at least one mutation with VAF above 0.05% was defined as ctDNA positive.

### Statistical analysis

The degree of linearity between K4C results and reference standards or benchmarked methods was assessed using R^2^ values derived from the “lm” linear model method within the R v4.3.0 Stats package. All graphical representations were generated using Rstudio server version 2022.07.02 (Rstudio Team, USA).

## Results

### Assay workflow

The K4C curated gene panel of 473 cancer-relevant genes included the entire exons, selected promoter and intron regions, with the total genomic size of 1.7 Mb (Table S3). FFPE and blood samples were required for the analysis (Figure 1). Genomic DNA from tumor and matching WBC samples were sequenced to profile all somatic and germline variants in 473 genes. The results could inform physicians of (1) actionable and resistant somatic mutations predictive of response to FDA-approved targeted therapies; (2) TMB and MSI status to guide immunotherapy decision, (3) pathogenic or likely pathogenic germline variants for genetic risk assessment. For selected 6 types of cancer, all tumor-derived mutations were further ranked by our proprietary algorithm and the top mutations were used to detect ctDNA presence in plasma using a bespoke mPCR assay. This result was used to (4) detect MRD and monitor treatment response for cancer patients.

**Figure 1.**
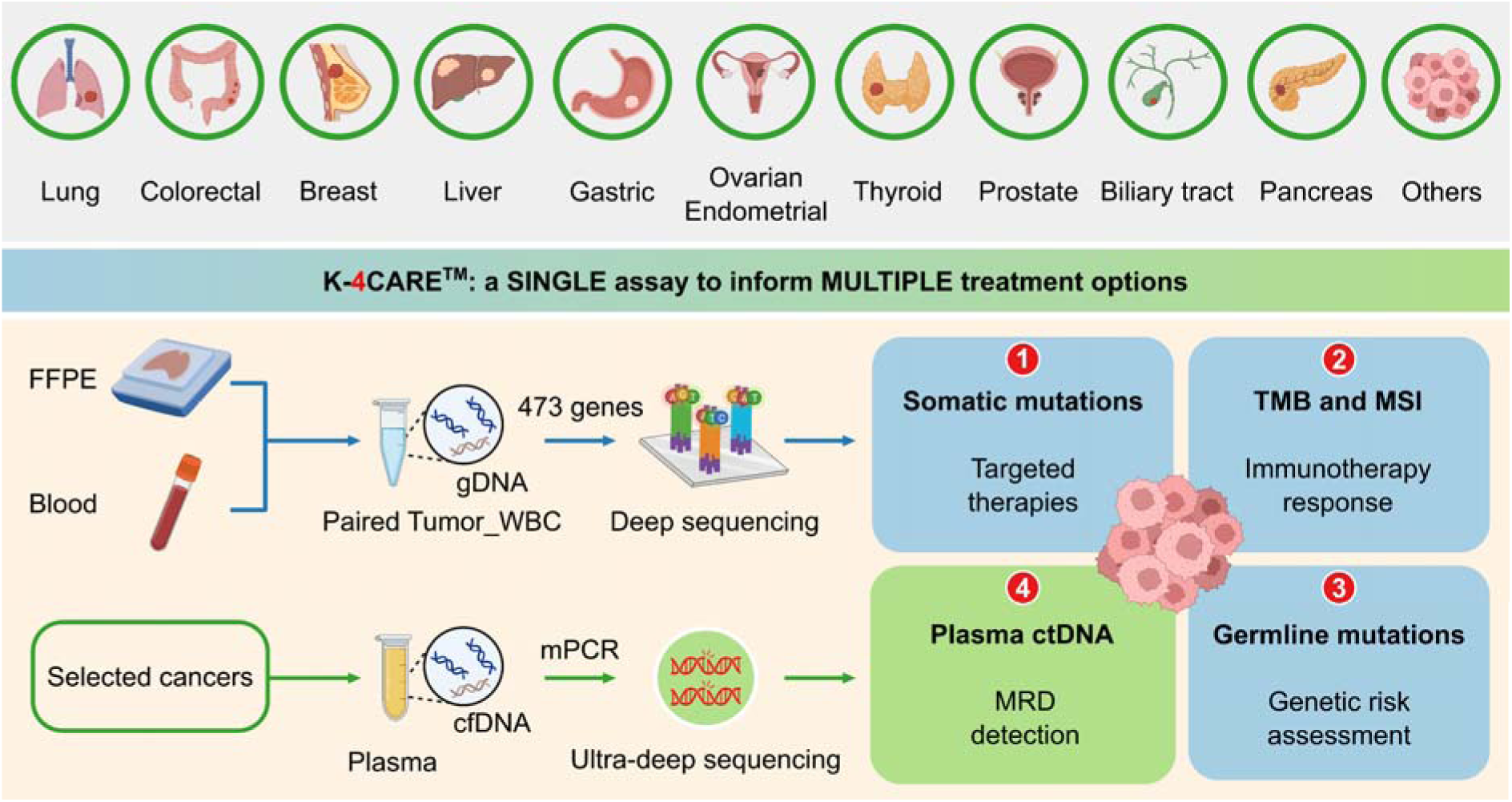
The K-4CARE^TM^ workflow. Formalin-fixed paraffin-embedded tumor tissue (FFPE) and peripheral blood sample of patients diagnosed with one of the 10 common types of cancer were collected. Genomic DNA (gDNA) from paired FFPE and white blood cell (WBC) samples of the same patient were hybridized with a panel of 473 cancer-relevant genes and subjected to deep sequencing (>150X) to identify somatic and germline mutations, tumor mutational burden (TMB) and microsatellite instability (MSI). For selected types of cancer (lung, colorectal, breast, liver, gastric and ovarian), cell-free DNA (cfDNA) was extracted from the plasma. Top ranked somatic mutations derived from the tumor were used in a bespoke multiplex PCR (mPCR) reaction followed by ultra-deep sequencing (100,000X) for detection of circulating-tumor DNA (ctDNA).

Analytical validation was first performed using reference standards to determine the accuracy and reproducibility of the K4C assay. Clinical samples from more than 10 types of cancer were then used to further evaluate the performance of the assay, especially when benchmarked against other orthogonal methods. The overall performance of the K4C assay is summarized in Table 1.

**Table 1.** Overall performance of K-4CARE^TM^ assay.

|  |  | **Sensitivity** | **Specificity** |
| --- | --- | --- | --- |
| **Targeted therapies (FFPE)** | **Somatic variants (at VAF ≥ 5%) \*** |  |  |
|  | SNV | >99% | >99% |
|  | INDEL | >99% | >99% |
|  | Amplification | >99% | >99% |
|  | Fusion | 94% | >99% |
| **Hereditary cancers (WBC)** | **Germline variants** |  |  |
|  | SNV | >99% | >99% |
|  | INDEL | >99% | >99% |
| **Immunotherapy (FFPE)** | **MSI** | 94% | >99% |
|  | **TMB** | **Correlation R-squared** |  |
|  |  | 97% |  |
| **Minimal residual disease (Plasma – Selected cancers)** | **ctDNA** | **Limit of detection** |  |
|  |  | VAF ≥ 0.05% |  |

\* For somatic SNVs and INDELs, variants at $1\% \leq \text{VAF} < 5\%$ were detected with sensitivity 82%, specificity >99%.
Abbreviations: ctDNA: circulating tumor DNA, FFPE: formalin fixed paraffin-embedded tissue, Indel: insertion/deletion, MSI: microsatellite instability, SNV: single nucleotide variation, TMB: tumor mutational burden, VAF: variant allele frequency, WBC: white blood cells.

### Detection of somatic and germline variants

The reference standard, OncoSpan gDNA, had 228 verified variants (211 SNVs, 17 INDELs) within interrogated region of our K4C gene panel. Sensitivity of K4C assay was estimated from 11 replicates: 2-3 replicates/run for 4 independent runs. For both SNV and Indel detection, sensitivity was estimated to be >99% for VAF ≥ 5% (Table S4). There was high correlation (R^2^ > 0.90) between the expected VAFs and VAFs determined by the K4C assay (Figure 2A, Table S5). Specificity was established at >99% for somatic SNV and Indel detection using Tru-Q 0 standard (Table S6). Limit of Detection (LOD) of the assay was defined as the minimum VAF detectable in at least 95% of technical replicates using the same input material. Using the OncoSpan standard, LOD for SNVs and Indels was determined at 5% VAFs (Table S7). Variants at 1% ≤ VAF < 5% were detected with sensitivity of 82% and specificity of >99% (Table S6-7). The high repeatability and reproducibility of the assay using the OncoSpan standard was shown in Table S8.

**Figure 2.**
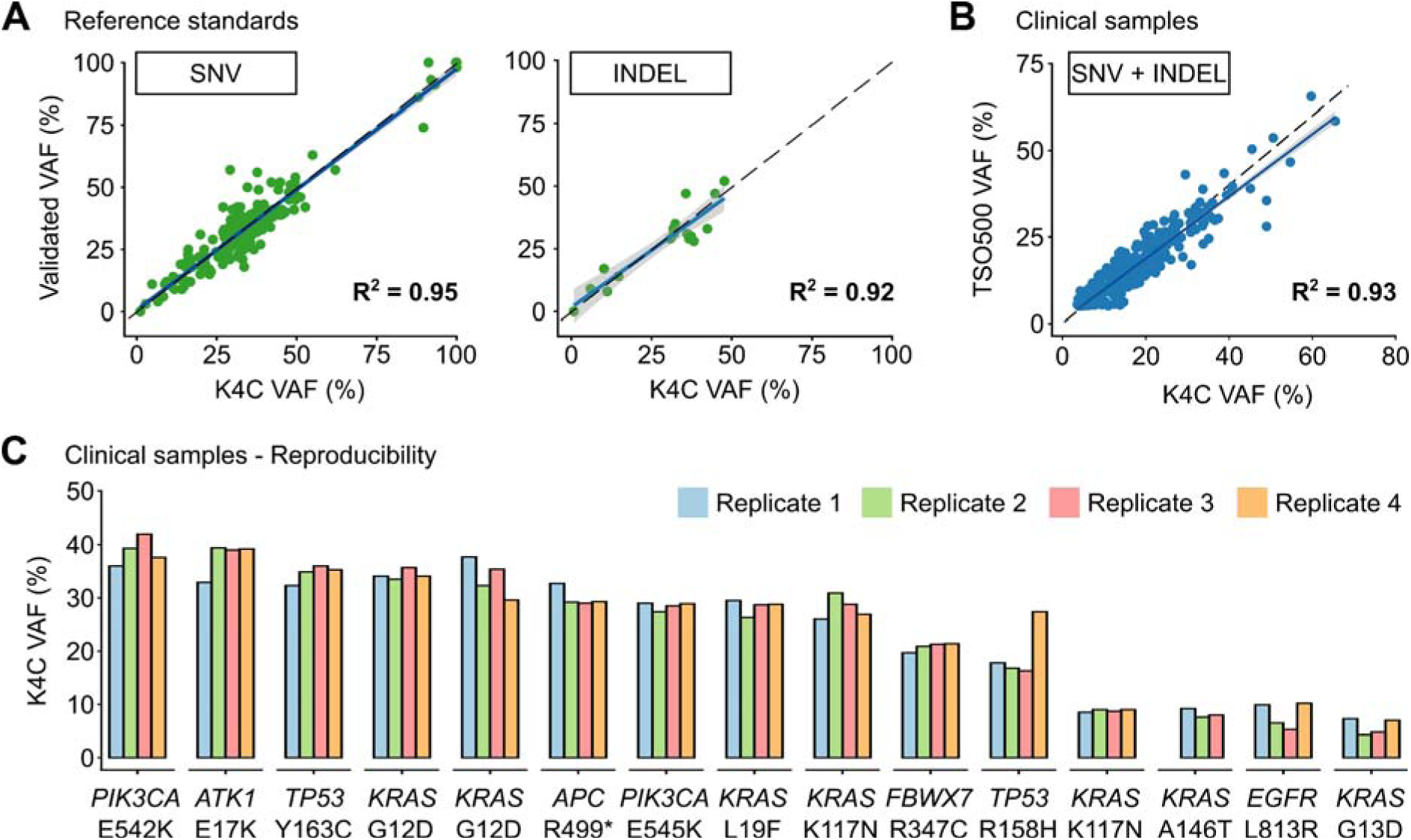
Performance of somatic SNV and Indel detection. **(A)** Validated variant allele frequencies (VAFs) of SNVs and Indels from Oncospan reference standards were compared with VAFs determined by the K-4CARE (K4C) assay. **(B)** For clinical samples, VAFs of SNVs and Indels identified by the K4C assay were compared with those detected by the commercial TruSight Oncology 500 (TSO500) assay (n=33 samples). The dashed line indicates x = y; the solid line is the regression line; and the gray area represents 95% confidence interval. **(C)** VAFs of SNVs and Indels of clinical samples measured as 4 independent replicates were reproducible (n=15 samples).

Next, we compared the performance to detect SNVs and Indels of the K4C assay with the TruSight Oncology 500 (TSO500) using 39 clinical samples. Concordance between the K4C and TSO500 assays to detect actionable SNV and Indel variants was 100%. The correlation of VAFs determined by two assays was 0.93 (Figure 2B). The high reproducibility of VAFs measured by the K4C assay was also demonstrated in clinical samples measured in 4 independent replicates (Figure 2C, Table S9).

For amplification, we used Breast CNV Mix and Lung & Brain CNV Mix standards comprising of 6 verified CNVs (*ERBB2, FGFR3, MYC, EGFR, MET, MYCN*) with 3 additional copies each. The K4C assay was able to identify amplification alterations in all six genes while it did not detect amplification in negative reference standards (Table S10). For gene fusion, Pan Cancer 6 Fusion standard that had six types of fusions: *TPM3-NTRK1*, *QKI-NTRK2*, *ETV6-NTRK3*, *EML4-ALK*, *CCDC6-RET*, and *SLC34A2-ROS1* was used. K4C assay detected 94% of fusion events in 3 independent replicates, while it did not detect any in negative reference standards (Table S11).

For germline variants, the K4C assay detected both SNV and Indel variants from the reference standard, BRCA germline I, with sensitivity and specificity of >99%; and with high correlation between expected VAFs and VAFs determined by the K4C assay (Table S12).

### Tumor mutational burden (TMB)

K4C assay determined the TMB of SeraSeq® TMB-7 and -13 standards as 7.7 and 14.2 respectively, which were within the recommended range from the manufacturer. Since these standards have a tumor fraction of 100%, which rarely applies to clinical samples, we next evaluated the stability of TMB values at lower tumor fractions and different sequencing performance. Fastq data of tumor standards were mixed with WBC standards at the ratios of 3:1, 1:1, 2:3, 3:7, 1:4, and 1:9 to create simulated tumor fractions ranging from 75% to 10%; followed by random read sampling to vary median depth and percentage coverage at 100X across different tumor fractions. We found that TMB values remained stable when tumor fraction was at least 30%, mean depth of at least 150X, and a percent coverage at 100X exceeding or equal to 75% (Figure 3A).

**Figure 3.**
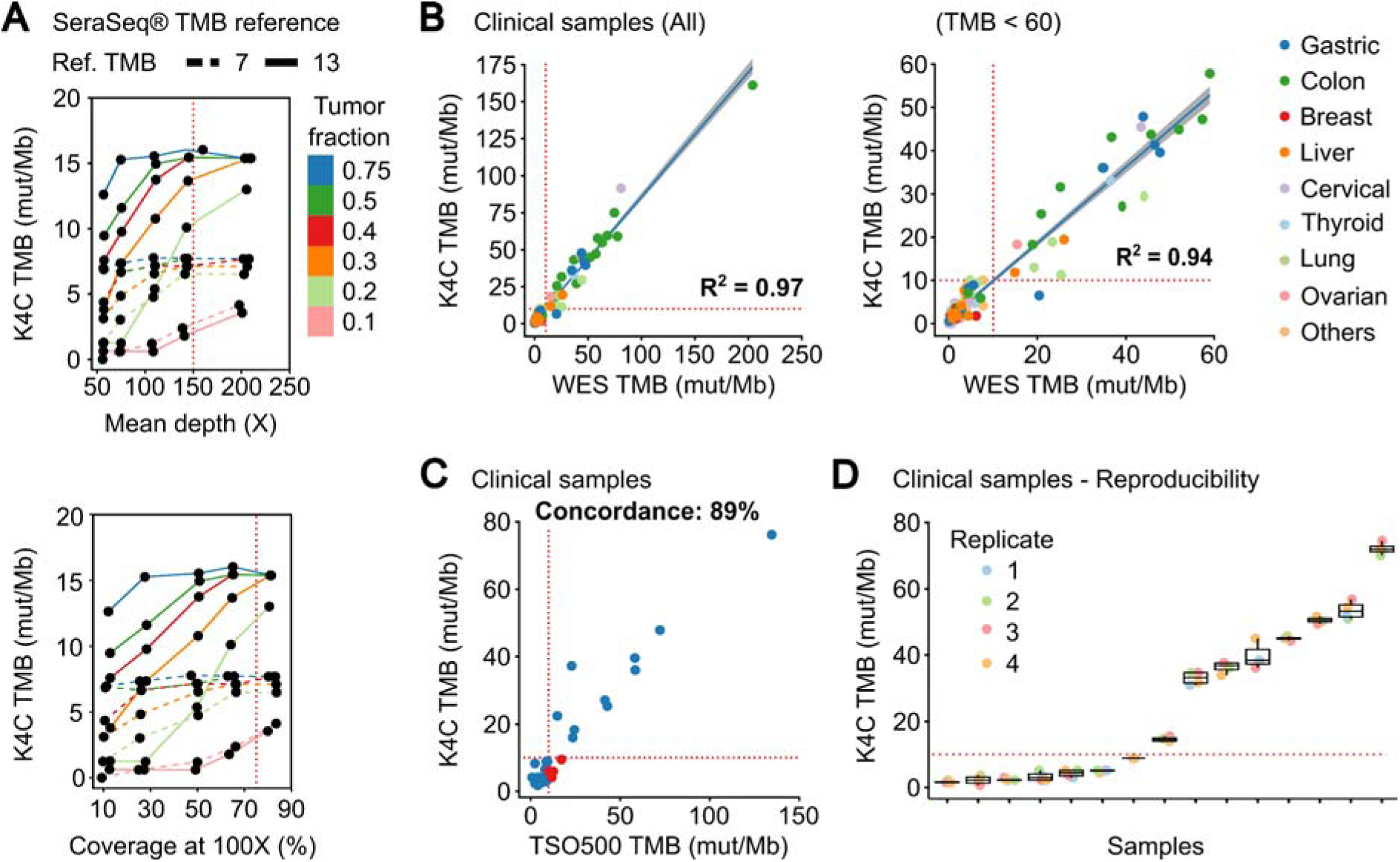
Performance of TMB measurement. **(A)** TMB values of the SeraSeq® TMB reference standards were measured by K4C assay when the following parameters varied: tumor fraction, mean depth and percent coverage at 100X. **(B)** Correlation of TMB scores determined using the K4C gene panel to those determined by whole exome sequencing (WES) in all clinical samples (n=127 samples), or samples with TMB < 60 (n=121 samples). **(C)** Concordance of TMB in clinical samples as determined by the K4C assay and the commercial TruSight Oncology 500 (TSO500) assay (n=35 samples). **(D)** TMB scores of clinical samples measured as 4 independent replicates by the K4C assay were reproducible (n=15 samples).

We then determined TMB values of 127 clinical samples using either K4C targeted panel or WES, which is the gold standard for TMB measurement. The correlation co-efficiency of TMB values between the 2 methods was 0.97 for all clinical samples, and 0.94 for samples with TMB < 60 (Figure 3B, Table S13). When benchmarked against the TSO500 assay using 39 clinical samples, the concordance to determine TMB status of the K4C assay was 89% (Figure 3C, Table S14). TMB scores were highly reproducible in clinical samples analyzed in 4 independent runs (Figure 3D).

### Microsatellite instability (MSI)

MSI status was first determined for 158 colorectal and gastric cancers, using PCR and fragment analysis for 5 microsatellite markers recommended by the National Cancer Institute [20]. K4C assay was performed for randomly selected 31 samples, including both MSI-H and MSS samples. The concordance of K4C assay to determine MSI status was 97% compared to the PCR method (Figure 4A, Table S15). When benchmarked against the TSO500 assay using 39 clinical samples, the concordance between K4C and TSO500 for MSI measurement was 98% (Figure 4B, Table S14). MSI scores determined by the K4C assay were highly reproducible in clinical samples analyzed in 4 independent runs (Figure 4C).

**Figure 4.**
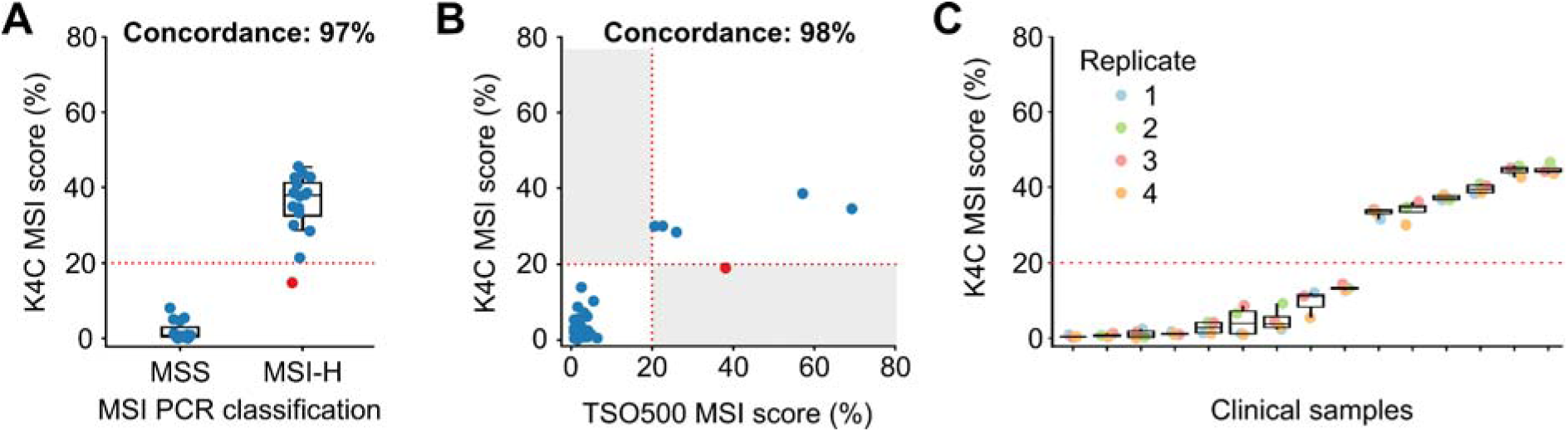
Performance of MSI measurement. **(A)** Concordance of MSI status determined by the K4C assay and by PCR-fragment analysis using gastric and colorectal cancers (n=31 samples). **(B)** Concordance of MSI status determined by the K4C assay and the commercial TruSight Oncology 500 (TSO500) assay for clinical samples (n=39 samples). **(C)** MSI scores of clinical samples measured as 4 independent replicates by the K4C assay were reproducible (n=15 samples).

### Clinical utilization

In order to illustrate the clinical usefulness of the K4C assay, we identified actionable alterations predictive of treatment response to FDA-approved drugs (OncoKB level 1 and 2 only). The percentage of samples with at least 1 actionable mutation was 68.4%, 57.1%, 50%, 47.1%, 9.1% and 7.1% in thyroid, breast, lung, CRC, ovarian and cervical cancers, respectively (Figure 5A, Table S16). The clinical evaluation of TMB showed that TMB score was highest in CRC, gastric and lung cancers (Figure 5B). For MSI, the prevalence of MSI-H was 16.9%, 9.1%, 5.3% and 3.4% in CRC, ovarian, thyroid and gastric cancers (Figure 5C). We also examined the relationship between TMB and MSI status in clinical samples (Table S17). Majority of the samples with concurrent TMB-H and MSI-H were CRC and gastric cancer. Several samples had TMB-H but MSS status, including lung and liver cancers (Figure 5D).

**Figure 5.**
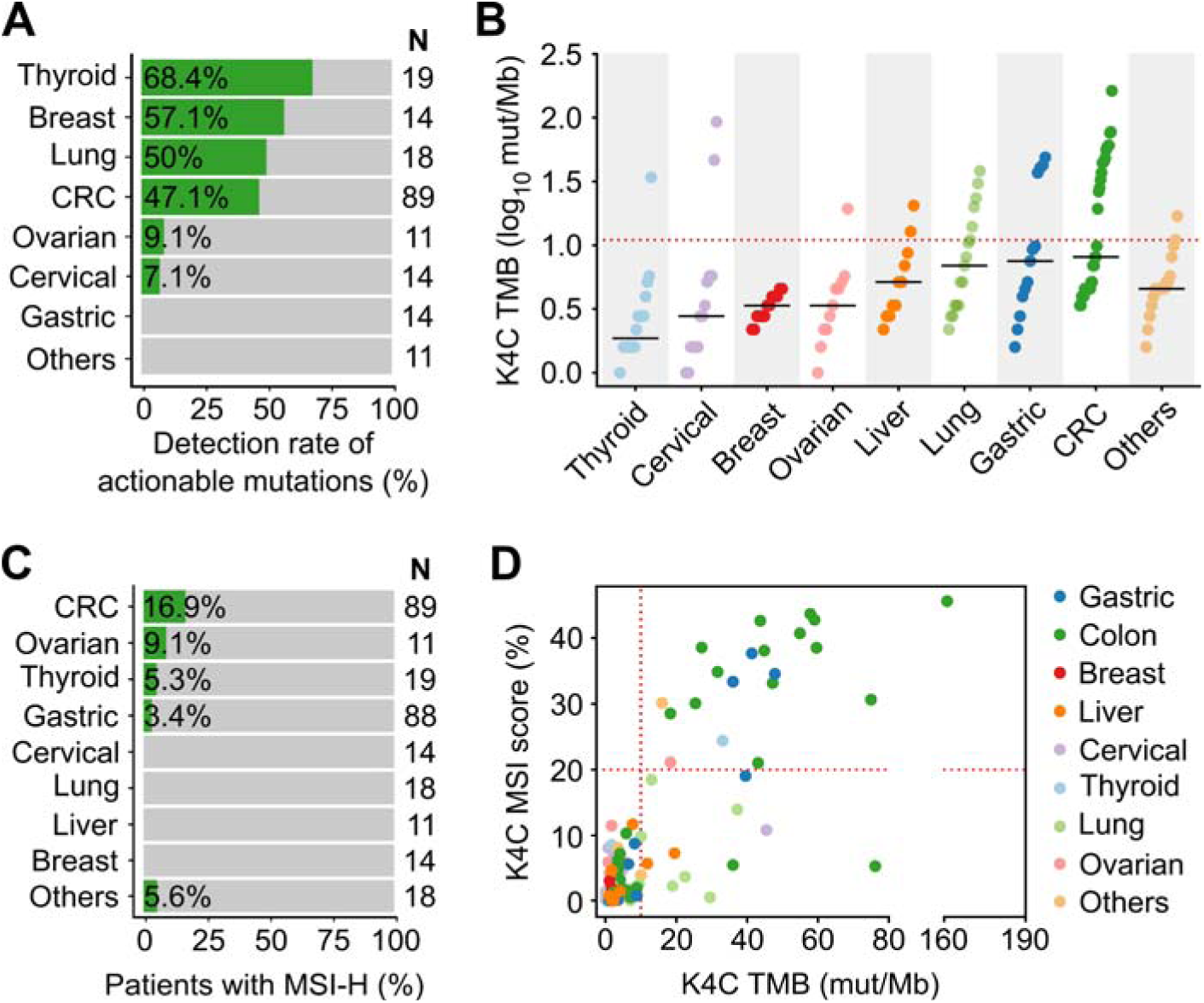
Clinical validation of the K-4CARE^TM^ assay. **(A)** Detection rate of actionable mutations that have FDA-approved drugs for different cancer types (n=190 samples). **(B)** TMB values (n=150 samples) and **(C)** MSI status (n=282 samples) across various cancer types. **(D)** Correlation of MSI and TMB in different cancers (n=150 samples).

### Detection of plasma ctDNA

In 40 patients with either CRC or breast cancer, the total number of tumor-specific mutations detected were 4257 variants. Using in-house ranking algorithm, we selected the top 2-5 variants unique for each patient and amplified them in bespoke mPCR assays to detect ctDNA in the plasma. Out of total 127 variants selected from tumors, 100 variants (78.7%) were detected in plasma samples (Figure 6A). There was no correlation between VAF of a variant in FFPE tissue and its VAF or detection status in the plasma (Figure 6B). When the same set of mutations were used to monitor ctDNA dynamics in longitudinal plasma samples, we could detect MRD and molecular relapse ahead of clinical diagnosis. The 2 case studies were presented to illustrate the clinical utility of the test. First, patient K4C012 was diagnosed with stage I, triple-negative breast cancer. ctDNA was detected in the pre-operative plasma, which was cleared after surgery but returned positive after adjuvant chemoradiotherapy (CRT). Clinical examination only confirmed cancer relapse 10 months after ctDNA was found positive (Figure 6C). Second, patient K4C077 was diagnosed with T4aN2M0 colon cancer. After surgery, VAF of ctDNA drastically declined but the patient remained ctDNA-postive, indicating presence of residual cancer cells. Despite CRT, he was diagnosed with liver metastasis 10.5 months later. Levels of carcinoembryonic antigen (CEA), the current biomarker used to monitor CRC patients, were normal both before and after surgery, and only rised at 6 months after ctDNA was found positive (Figure 6D).

**Figure 6.**
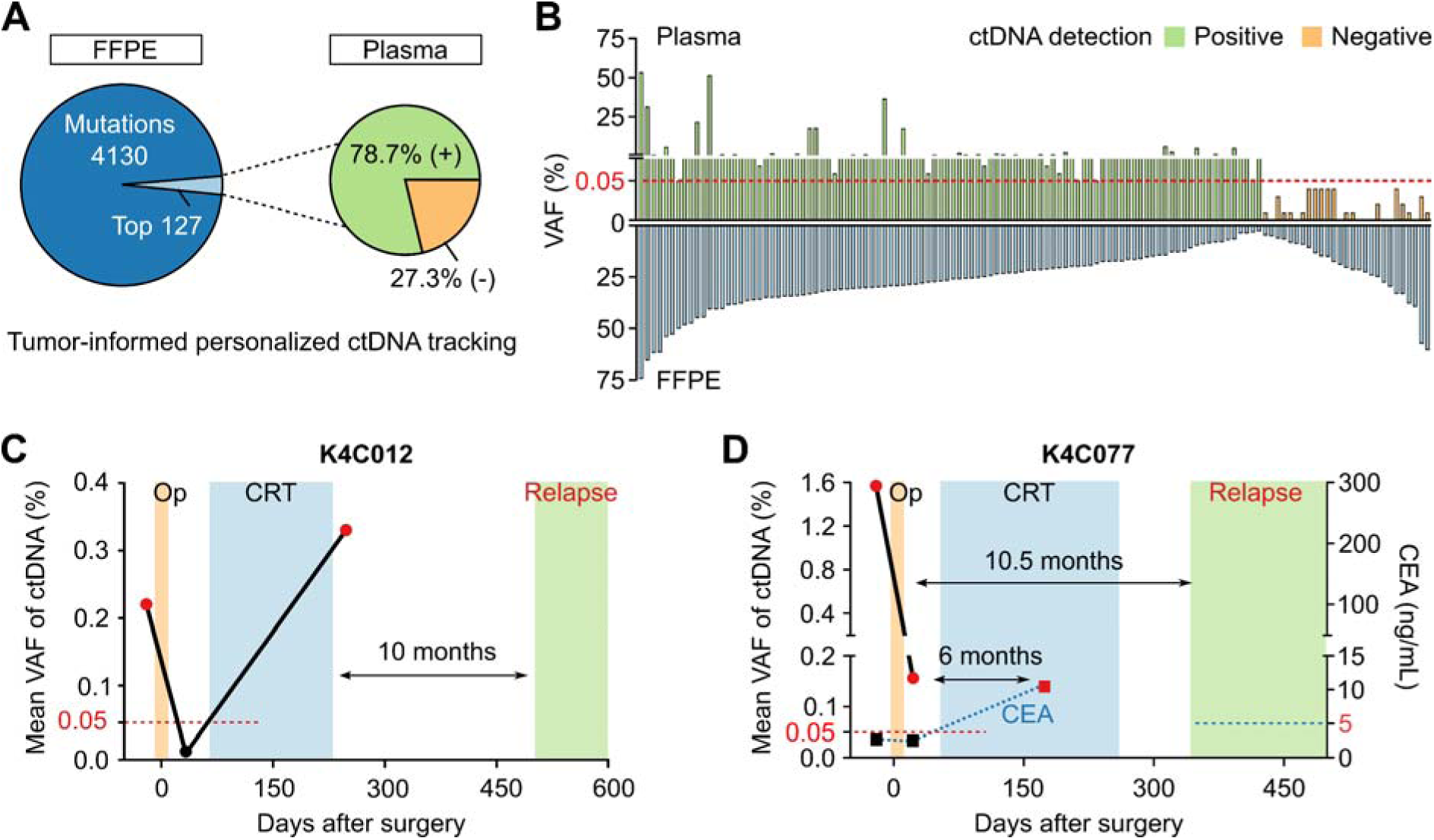
Plasma ctDNA tracking to monitor treatment response. **(A)** In 40 patients with either breast cancer or CRC, in-house ranking algorithm was used to select top variants for each patient to track ctDNA presence. 100 out of 127 variants (78.7%) were detected in the plasma. **(B)** No correlation between VAF of a variant in FFPE tissue and its VAF or detection status in plasma. **(C-D)** Case study showing longitudinal ctDNA monitoring and clinical status of 2 patients diagnosed with breast cancer (K4C012) and CRC (K4C077). Op: operation, CRT: chemoradiotherapy, CEA: carcinoembryonic antigen.

## Discussion

In this study, we presented the analytical validation and clinical utilization of K-4CARE^TM^, a tumor-based CGP assay that enables the detection of somatic variants, germline variants, TMB and MSI measurement, as well as ctDNA monitoring, all within a single platform. Performance metrics of the assay have been rigorously established and validated using both reference standards and orthogonal methods.

Firstly, the K4C assay could detect 4 types of somatic variants with high sensitivity and specificity, using tumor DNA only. Unlike other CGP assays that used RNA to detect gene rearrangement [21; 22; 23], we designed probes to target selected intron regions of commonly fused genes, and DNA libraries were used to hybridize with these probes. Our bioinformatic pipeline to detect gene fusion was robust and similar to those developed in previous studies [24; 25; 26]. Since this eliminates the necessity to process tumor RNA, it expedites the result and makes the assay more cost-effective and feasible, particularly for developing countries like Vietnam where the quality of FFPE-derived RNA is often not sufficient for NGS (in-house data). Furthermore, we demonstrated clinical usefulness of our assay to identify at least 1 actionable mutations (OncoKB level 1-2) in 38.9% of the clinical samples (74/190). This rate seemed to be higher than other CGP assays applied in other cohorts, such as 12.0% in Ng *et al* [21], 10.7% in Aoyagi *et al* [27] and 23.7% in Karol *et al* [28]. It was likely attributed to the distribution of samples across different cancer types in each cohort. Our analysis included more CRC (46.8%) and thyroid cancer (10.0%), both of which have high prevalence of actionable mutations, compared to other cohorts. Particularly for papillary thyroid cancer, we identified the prevalence of *BRAF* V600E mutations in 68.4% of the samples, which was at the high end of the reported range (32-65%) [29]. Since the sample size in each cancer type was small, this result was solely used to illustrate the clinical utilization of the assay, rather than providing representative mutational spectrum or true level of actionability. More comprehensive analysis of actionable mutation profiles in substantially larger cohorts of lung, breast and colorectal cancer was reported in our previous studies [5; 9; 10].

For TMB measurement, we included non-synonymous mutations in a genomic region of approximately 1.7 Mb, larger than the recommended 1 Mb to ensure accuracy and stability of TMB [2; 30]. The validation process has been aligned with current standards and previous CGP assays [21; 22; 23; 31], showing high correlation between K4C panel and the gold standard WES (R^2^ = 0.97). The concordance of TMB measurement between K4C (tumor-normal) and TSO500 (tumor-only) assays was 89%, slightly lower than the concordance among tumor-only panels (>90%) [30]. We observed that all non-concordant samples had higher TSO500 TMB compared to the K4C value, which was also reported for a different tumor-normal CGP assay [23]. Such discrepancy was likely due to the absence of matched normal sample and the inclusion of synonymous mutations in the TMB calculation algorithm of the TSO500 assay [23]. In addition, among our clinical samples, cancer types with the highest TMB scores were CRC, gastric and lung cancers. This result was consistent with previous studies [23; 32; 33] but different from the Ng *et al* study reporting the highest TMB in liver and breast cancers in an Asian cohort [21].

For MSI analysis, IHC and PCR-based assay examining 5 mononucleotide repeat markers remain the first choices in cancers belonging to the spectrum of Lynch syndrome such as CRC and endometrial cancer [34]. MSI evaluation by NGS is also recommended as an option for these cancers for the additional benefit of examining TMB, another biomarker for ICIs [34]. For all cancers not related to Lynch syndrome, NGS is the method of choice to determine MSI as the reliability of IHC and the 5 markers by PCR has not been proven [34]. In our analysis using CRC and gastric cancer samples, we demonstrated excellent concordance of MSI measured by PCR for 5 repeat markers and NGS examining ∼800 repeat loci, indicating the reliability and accuracy of the NGS technique. The prevalence of MSI-H was 16.9% in CRC, similar to the rate reported previously [35; 36]. The prevalence of MSI-H in our gastric cancer samples (3.4%) was lower than that in Bonneville *et al* (19.9%) [35] but comparable to other large cohorts (2.5-3.4%) [36; 37]. The surprisingly high incidence of MSI-H in ovarian and thyroid cancers in our cohort needs to be further investigated with a larger sample size.

Since PD-L1, MSI and TMB have been approved as independent biomarkers for ICIs, the relationship among these three has been extensively examined in previous studies. PD-L1 was found to have low concordance with TMB and MSI, while the relationship between TMB and MSI varied significantly by cancer type [33; 34]. The highest concordance of TMB and MSI was found in CRC and gastric cancer, and almost none in lung cancer [33; 34]. This completely agreed with our observation in this study. Moreover, similar to those studies, we also found TMB-H could occur in MSS samples across different cancer types, while the incidence of MSI-H in TMB-L samples was rare. This again emphasizes the importance of comprehensive testing for multiple biomarkers to maximize the treatment opportunity for patients. It should also be noted that the interpretation of these biomarkers has to be cancer type-specific, as the cut-off for TMB-H at 10 mut/Mb, for example, has been shown to be only applicable for lung carcinoma [38].

Lastly, a unique application of the K4C assay is the ability to use personalized mutations derived from tumor to track for the presence of ctDNA in plasma. The analytical validation of the mPCR technique was already elaborated in our previous studies [9; 10]. At the LOD of 0.05%, the assay robustly and accurately identified ctDNA in the plasma using mutations ranked by our in-house algorithm. The prognostic and predictive value of ctDNA in routine monitoring of cancer patients during and after treatment have been consistently demonstrated in several clinical trials [8]. Specifically for ICIs, ctDNA clearance after 2 cycles of treatment was shown as a strong indicator of long-lasting response in multiple solid tumors [39; 40].

Taken together, the data presented in this study highlights the accuracy and reliability of the K4C assay in providing comprehensive genomic alterations and signatures to inform targeted and immunotherapies for cancer patients. The assay could also detect ctDNA in the plasma, that can later be used to routinely monitor treatment response and detect residual cancer.

## Ethical statement

The use of FFPE and WBC samples at the Medical Genetics Institute was approved by the institutional ethics committee of the University of Medicine and Pharmacy, Ho Chi Minh city (approval number 164/HDDD). The use of plasma samples for ctDNA tracking was under the studies approved by the ethics committee of the University of Medicine and Pharmacy, Ho Chi Minh city (approval number 14/GCN-HDDD for colorectal cancer, 300/HDDD for breast cancer). All genomic data were de-identified and aggregated for the genetic analysis.

## Author contributions

Thien-Phuc Hoang Nguyen, Tien Anh Nguyen, Hoa Giang performed bioinformatics and statistical analysis. Nam HB Tran, Van-Anh Nguyen Hoang, Vu-Uyen Tran, Yen Nhi Nguyen, Anh Tuan Nguyen, Hong Thuy Thi Dao, Cam Tu Nguyen Thi processed standard and clinical samples for sequencing. Thanh Thuy Do Thi, Duy Sinh Nguyen, Hoai-Nghia Nguyen performed clinical analysis. Thien-Phuc Hoang Nguyen, Lan N Tu designed experiments, analyzed data and wrote the manuscript.

## Acknowledgements

We thank Dr Hoa Phan, Medical Genetics Institute, Vietnam for assisting in manuscript preparation.

## Data availability statement

The data presented in the study are deposited in the BioProject repository, accession number PRJNA1035299.

